# Eligibility Without Equity: Rethinking Age-Based Adult Vaccine Policies

**DOI:** 10.64898/2026.02.17.26346473

**Authors:** Mohammed Sherif Amin, Brendan Collins, Catherine Beavis, Jennifer Sigafoos, Neil French, Daniel Hungerford

## Abstract

Embedding equity into vaccine eligibility is essential for reducing health inequalities. Yet, adult vaccine eligibility in most European countries is primarily based on fixed age thresholds, prioritising cost-effectiveness. This approach risks excluding the most vulnerable populations living in deprived communities with poorer health and shorter survival into older age. Extending eligibility based on clinical risk partially addresses this gap. Higher rates of underdiagnosis and delayed diagnosis in deprived populations limit the fairness of this approach, however, with the status quo of adult vaccine eligibility criteria likely doing active harm. In this perspective, we demonstrate the extent of this inequity in England. For example, the average male living in Hyde Park in the northern city of Leeds dies 9.5 years too early to ever receive the RSV vaccine offer at age 75. Meanwhile, a male living in Hyde Park, London, lives much longer and may receive the benefits of the RSV vaccine for 10 years or more. Drawing on lessons from the COVID-19 pandemic, we propose further evaluation of alternative eligibility models that incorporate local place-based disadvantage, which will inherently account for life expectancy and deprivation levels. These models will ensure earlier access to vaccines for communities with the greatest need and improve health equity without overwhelming health systems.

## Inequity in the age-based vaccine offer for adults

Vaccines are our best defence against infectious diseases and are among the most cost-effective public health interventions [1]. Vaccination programmes often aim to achieve equity through a universal offer. For childhood vaccines, this is typically achieved through a national schedule where every child, regardless of demographics or where they live, is eligible for the same vaccine offer.

Adult vaccination programmes operate differently and are inherently more complex. They must account for heterogeneous risk profiles across life course, competing health priorities, and the practical challenges of programme delivery at scale. As a result, National Immunisation Technical Advisory Groups (NITAGs) must balance complexity with parsimony in their recommendations.

Within this framework, eligibility for adult vaccines in most European countries, including the UK, is largely defined by cost-effectiveness-guided fixed age thresholds that serve as proxies for senescence, frailty, and population-level disease risk [2]. However, cost-effectiveness analyses are inherently utilitarian, prioritising aggregate population health gains rather than the distribution of those gains across social groups. Individuals who are less likely to survive into very old age, or whose health gains are more costly to achieve, may accrue fewer benefits, even when their underlying vulnerability is greater.

When eligibility is set at fixed chronological age thresholds, particularly at advanced ages, these dynamics intersect with socially patterned differences in survival. A smaller proportion of people living in more deprived areas ever reach the cutoff at which vaccination is offered, resulting in social injustice and an inequitable vaccine eligibility.

Recent policy documents, including the 2025 UK Health Security Agency’s Immunisation Equity Strategy [3], and the European Centre for Disease Prevention and Control (ECDC) annual influenza vaccination report [4] highlighted persistent inequalities in infectious disease outcomes in older adults and called for a systematic approach to prevention in later life, acknowledging that uniform models of delivery do not meet the needs of all communities. Yet, much of the discussion focuses on equality of access among those who are already eligible, rather than the deeper issue of who actually gets the chance to benefit in the first place (i.e., equity of eligibility). In some communities, people are dying before they ever become eligible for the vaccines designed to protect them.

Taking the influenza (Flu) vaccine as an example, older adults in the UK [5], along with other European countries [6], are offered free annual flu vaccines from age 65 years. The public health benefit will be approximately tied to the number of years a person remains eligible. An individual who lives for 20 years after crossing the vaccine eligibility threshold will enjoy more years of protection (assuming vaccine take up) than someone who dies early at 70 due to inequalities in the social determinants of health and the very viruses that these vaccines protect against. In this case, a longer life simply means a greater benefit and widening health inequalities.

It is also important to acknowledge that most adult vaccines incorporate additional criteria for eligibility in some circumstances, including clinical susceptibility to the vaccine preventable disease. This means that younger adults from deprived areas may qualify for vaccination if they have certain long-term conditions like asthma, COPD, cardiovascular disease, or diabetes. Adults in the UK under 65 with these conditions are offered the annual flu vaccination, and similar policies exist for COVID-19 and pneumococcal vaccines [7]. Yet this dual eligibility approach does not necessarily reduce inequalities in outcomes, as not all outcome inequalities are mediated by the presence of eligible diagnosed conditions. Many are mediated by social factors such as occupation, income, housing and environmental exposures [8], and these determinants are not currently being considered in the adult vaccination programmes’ eligibility criteria.

Furthermore, residents of deprived areas often face delays or underdiagnosis of long-term conditions [9,10]. According to the 2025 UK Health Inequalities in Health Protection report [11], emergency hospital admissions for infectious diseases are almost double in areas from the most deprived quintile of England compared to the least deprived, with an estimated 260,000 additional admissions of respiratory infections alone attributable to deprivation. Consequently, even when the offer is extended to include pre-existing clinical diagnoses, those living in the poorest environments remain the least likely to receive timely protection.

Inequity in offer is even more pronounced for vaccines given later in life, such as the newly introduced respiratory syncytial virus (RSV) vaccine. The RSV vaccine was initially offered in the UK for adults aged 75-79 and in July 2025, the Joint Committee on Vaccination and Immunisation (JCVI), the UK’s NITAG, extended the eligibility to include individuals aged 80 and over [12], aligning with other countries as Germany [13] and Sweden [14].

## Life expectancy and years of eligibility

To illustrate the scale of this inequity, we present a visual comparison (Figure 1) showing how vaccine eligibility can vary dramatically by where someone lives. Using England’s life expectancy at birth (2019-2023) [15], we estimated the number of years people would remain eligible for the seasonal influenza vaccine (from age 65) and RSV (from age 75), stratified by sex. In many parts of the North of England and the Midlands, areas with high levels of deprivation, individuals can expect as few as zero to five years of eligibility for flu vaccine. Similarly, for some areas RSV vaccine is offered too late to ever be received, as average life expectancy falls below 75. For instance, in the Hyde Park area of Leeds, a city in the North of England, males have among the lowest life expectancy figures, at 65.5 years, which corresponds to dying on average 9.5 years before becoming eligible for the RSV vaccine and 0.5 years of flu vaccine eligibility. Meanwhile, residents of the more affluent Hyde Park area in London are likely to benefit from a substantially longer eligibility period, reaching up to 10.8 and 20.8 years for the RSV and flu vaccines, respectively. We can also observe that males are likely to have fewer years of vaccine eligibility in both cases than females.

**Figure 1.**
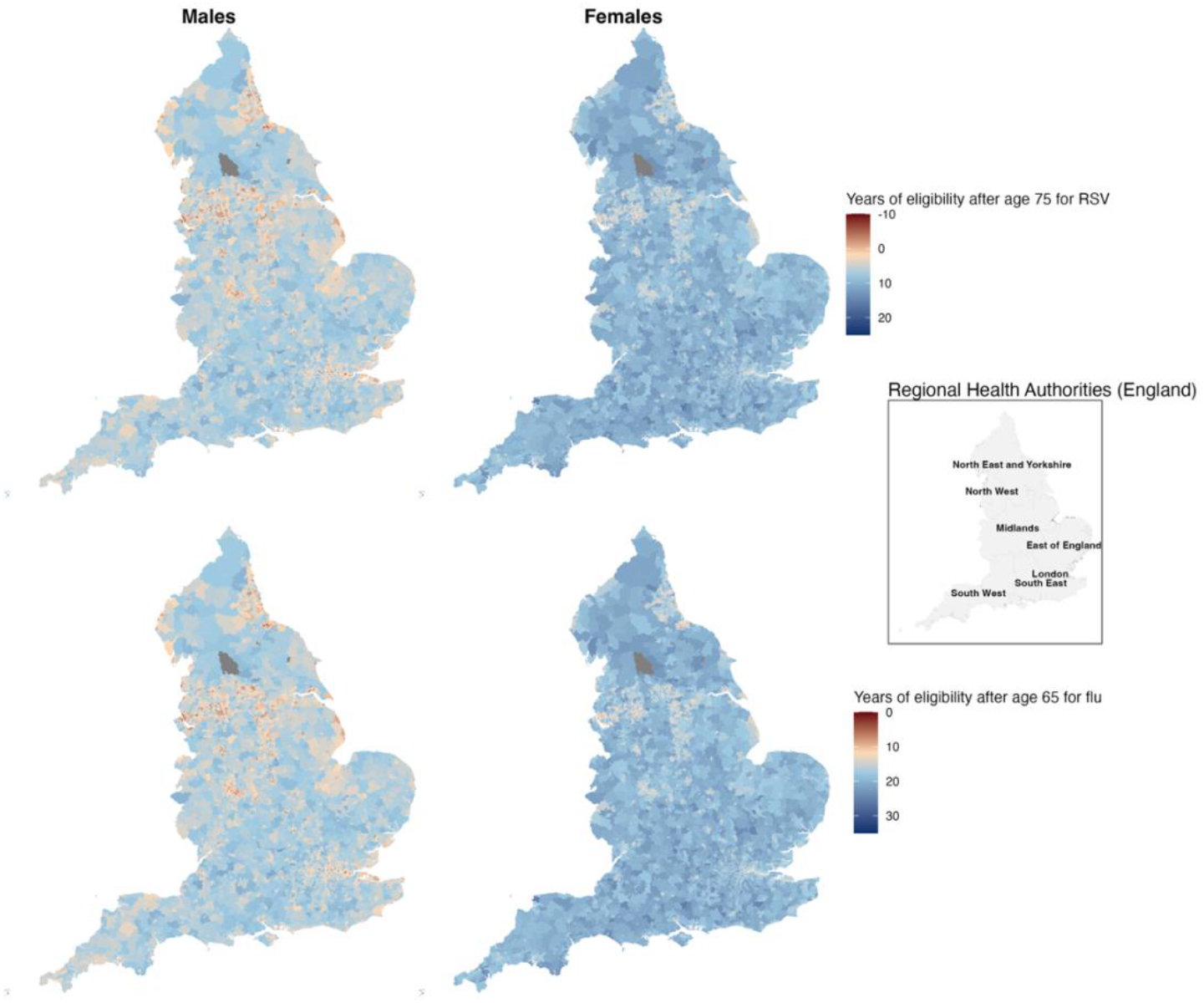
Estimated years of eligibility after age 75 (RSV) and 65 (flu) by sex and average life expectancy in England. Maps created by comparing the age-based vaccine eligibility criteria (75+ for RSV and 65+ for flu) to the average life expectancy (2019-2023) in each small area neighbourhood in England. Dark red indicates a shorter period of potential eligibility.

As at-birth life expectancy may be influenced by mortality occurring earlier in life, we further examined this pattern using survival to vaccine eligibility ages from the 2018-2020 English life tables [16], stratified by socio-economic deprivation, measured using the 2019 English Index of Multiple Deprivation [17]. Clear socioeconomic gradients in survival are evident, with differences more pronounced from mid-adulthood (around age 45 years) and widening progressively into older age, resulting in large absolute differences in eligibility (Figure 2). Around 77 in every 100 males living in the most deprived decile survive to the age at which influenza vaccination is offered (65-69 years), compared with approximately 92 in every 100 in the least deprived decile. At older ages, the gap widens further: only around 55 in every 100 males in the most deprived decile survive to the RSV eligibility age (75-79 years), compared with more than 82 in every 100 in the least deprived decile.

**Figure 2.**
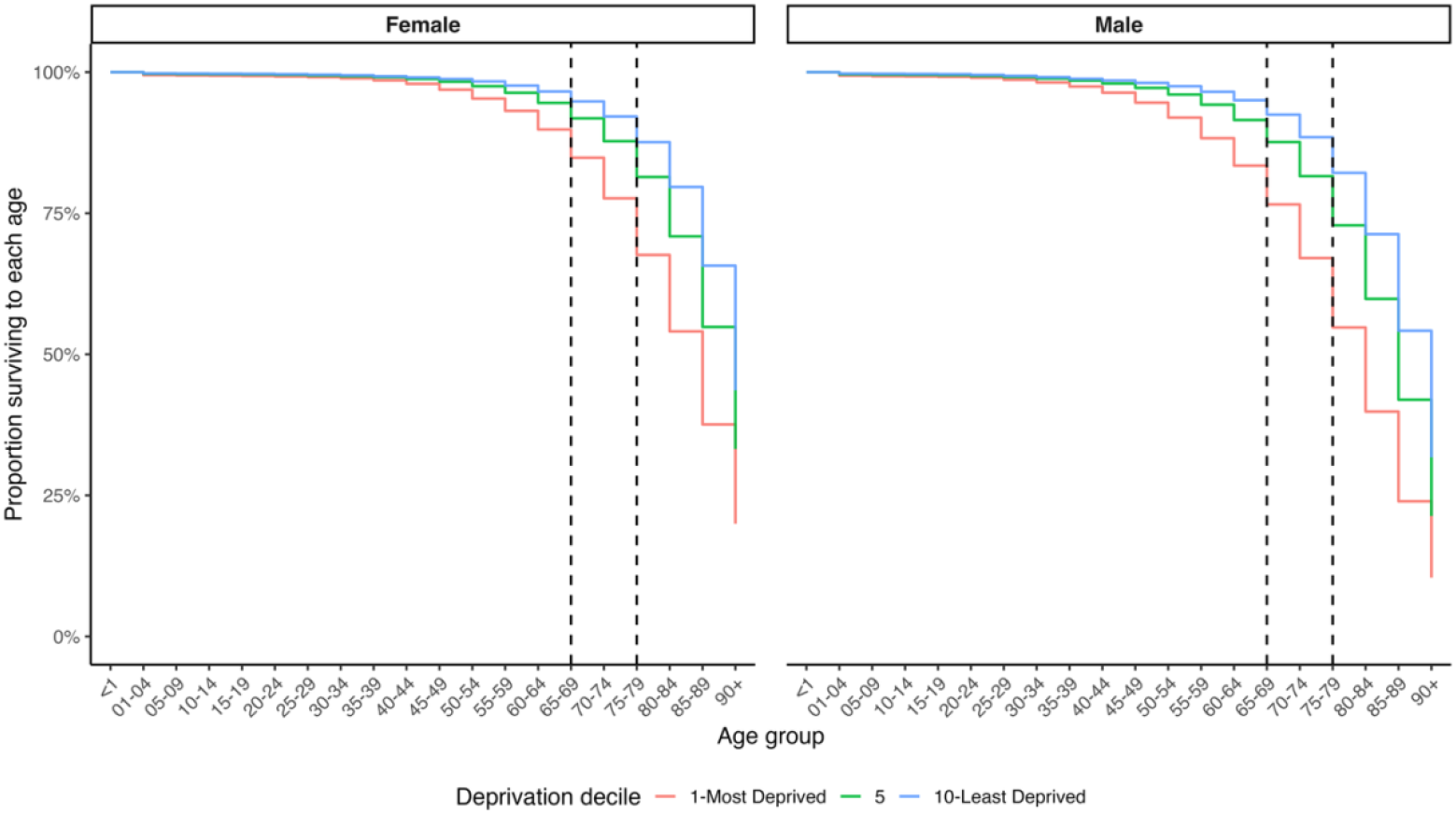
Survival curves by national deprivation decile in England, illustrating socially patterned differences in survival across the life course. Curves represent the proportion of a birth cohort surviving to each age group, based on life table data for England (2018-2020). Dashed vertical lines mark current age-based eligibility thresholds for seasonal influenza (65-69 years) and RSV vaccination (75-79 years). Data is open access through the UK Office for National Statistics website.

A comparable situation can be observed in other high-income European countries, though to a lesser extent. In Germany, male life expectancy between 2022 and 2024 in Sachsen-Anhalt, in East Germany, was 75.9 years, compared with 78.9 years in Hessen [18], a more affluent state in the west, which leads to the same dilemma of inequitable vaccine offer.

The reliance on chronological age highlights Tudor Hart’s ‘inverse care law’. People living in healthier, wealthier areas have greater vaccine access, while a high proportion of those most vulnerable to respiratory disease, often living in more deprived areas [19], are entirely excluded. This exclusion is not a matter of access or uptake, but of a policy design that conditions vaccine eligibility on age, without adequate consideration of social determinants of health, where someone’s location, and consequently socioeconomic position, is a predictor of whether they will live long enough to benefit from protection. This contrasts with the European Union’s [20] and England’s Healthy Ageing plans [21], which both emphasise reducing health inequalities among vulnerable groups by targeting social determinants of health.

## Alternative model to adult vaccine eligibility

It seems inadequate to determine vaccine eligibility solely based on crude age boundaries that do not account for existing health inequalities. The notion of optimising vaccine eligibility was previously proposed during the COVID-19 pandemic [22], although the primary motive at the time was to manage demand that exceeded limited vaccine supply. Subsequently, several approaches were trialled and demonstrated how health interventions could be better tailored to risk.

One of such approaches was the QCOVID algorithm [23] developed to estimate individuals’ risk of severe COVID-19 outcomes. This algorithm incorporated socioeconomic position with clinical risk factors to predict the likelihood of hospitalisation or death from COVID-19 infection. While QCOVID was primarily used for shielding advice, not vaccine eligibility, it raises the possibility of adapting an analogous algorithm for vaccine preventable diseases like influenza and RSV, integrating area or household socioeconomic factors, so that vaccines can be better targeted to the population at highest risk.

A similar example was the tiered local public health and social measures restriction system implemented in the UK as part of the COVID-19 response [24], where the stringency of control measures varied across local authorities (cities) and were continuously being updated based on real-time local infection rates.

Conceptually, both approaches share the same underlying principle; interventions were tailored to local risk instead of being applied uniformly to ensure protection was directed to areas with the greatest need. Although these represent a potential risk-based model for refining adult vaccine eligibility, implementing a continuously updating complex algorithm on a national scale might be operationally challenging. But this does not mean it should not be considered.

An alternative way would be to adopt a people and place-based model, given that socioeconomic inequalities, one of the main drivers of differences in life expectancy, are largely geographically patterned. Instead of a single universal age threshold, vaccine eligibility could be adjusted for both people level clinical risk and small-area life expectancy/survival, such that residents in more deprived neighbourhoods with lower average life expectancy become eligible earlier. For example, if the average life expectancy in a small area neighbourhood falls below a defined threshold, the age cut-off for vaccination could be lowered proportionally.

Importantly, the technical infrastructure required to support such an approach already exists within national immunisation systems (NHS). The Eligibility Data Product (EliD), currently being trialled within NHS England, integrates multiple data sources to determine vaccine eligibility [25]. Although EliD is currently based on age and clinical risk, it already incorporates place-based information for targeted signposting. Extending such systems to incorporate carefully defined place-based modifiers to age thresholds would therefore represent a policy choice rather than a technical barrier.

Vaccination eligibility frameworks that better reflect socially patterned differences in survival and vulnerability have the potential to address the inequity in current adult immunisation programmes. Whether such approaches would translate into improved health outcomes, represent good value for money, or be acceptable at scale remains an empirical question. This piece highlights the need for further work to evaluate the feasibility, effectiveness, and cost-effectiveness of alternative eligibility models that incorporate both individual-level risk and place-based disadvantage. As the European governments consider policies to extend working lives, developing and testing immunisation strategies that more equitably distribute opportunities for prevention across the life course should be a priority for future research and policy evaluation.

## Conclusion

Current vaccine eligibility policies that primarily rely on fixed age thresholds overlook critical social inequalities, excluding those most vulnerable due to shorter life expectancy. The result is missed opportunities to improve the quality of life and perpetuation of inequalities in healthy life expectancy. Using people- and place-based eligibility criteria, which inherently account for local life expectancy and levels of deprivation, could represent a pragmatic alternative for providing targeted protection for those at greatest risk. Embedding equity into the vaccine eligibility will help build public trust in government and medicine in underserved communities. Future research and policy should prioritise these approaches to build a fairer adult immunisation system.

## Contributions

DH, MSA, NF, and BC contributed to the conceptualisation of the idea. MSA carried out the formal analysis and wrote the original draft. All authors reviewed, edited and approved the final draft.

## Data availability and ethics statement

All the data used are publicly available on the Office for National statistics website (https://www.ons.gov.uk) and Fingertips platform provided by the UK Department of Health and Social Care (https://fingertips.phe.org.uk).

## Acknowledgements

None

